# Safety of Focused Ultrasound Neuromodulation in Humans with Temporal Lobe Epilepsy

**DOI:** 10.1101/2020.04.10.20060855

**Authors:** John M. Stern, Alexander S. Korb, Norman M. Spivak, Sergio Becerra, David Kronemyer, Négar Khanlou, Samuel D. Reyes, Martin M. Monti, Caroline Schnakers, Patricia Walshaw, Taylor P. Kuhn, Mark S. Cohen, William Yong, Itzhak Fried, Sheldon E. Jordan, Jerome Engel, Mark E. Schafer, Alexander Bystritsky

## Abstract

**Objective:** Low-Intensity Focused Ultrasound Pulsations (LIFUP) is a promising new potential neuromodulation tool. However, the safety of LIFUP neuromodulation has not yet been adequately assessed. Patients with refractory temporal lobe epilepsy electing to undergo an anterior temporal lobe resection present a unique opportunity to evaluate the safety and efficacy of LIFUP neuromodulation. Because the brain tissue in these patients will be removed, histological changes in tissue after LIFUP can be examined. Evidence of effective neuromodulation was assessed using functional MRI and EEG, while further potential safety concerns were assessed using neuropsychological testing.

**Methods:** EEG, functional MRI, and neuropsychology were assessed in six patients before and after focused ultrasound sonication of the temporal lobe at intensities up to 5760 mW/cm^2^. Using the BrainSonix Pulsar 1002, LIFUP was delivered under MR guidance, using the Siemens Magnetom 3T Prisma scanner. Neuropsychological changes were assessed using various batteries. EEG was recorded using the Electrical Geodesics EGI 256 channel system. Histological changes were assessed using hematoxylin and eosin staining, among others.

**Results:** LIFUP was not able to modulate the BOLD signal on fMRI in a reliable and consistent manner. The EEG data that was available did not demonstrate a change in activity after LIFUP in all but one subject. Likewise, the neuropsychology testing did not show any statistically significant changes in any test, except for a slight decrease in performance on the one test after LIFUP. Lastly, the histology did not reveal any detectable damage to the tissue, except for one subject for whom the histology findings were inconclusive.

**Significance:** The safety in the histology was the primary endpoint, and as such, longer exposure at the highest intensity levels will be administered moving forward.

**Key Points:**

- LIFUP is a novel brain stimulation technique with not yet fully established safety guidelines.
- LIFUP was administered to patients electing to undergo resective brain surgery.
- LIFUP does not appear to cause damage to tissue.
- Longer exposure times are needed to further show safety at these intensity levels.

## Introduction

Non-invasive, controllable, and reversible modulation of regional brain activity is a major interest in current neuroscience because current neuromodulation technologies are *either* invasive, or limited in their spatial resolution. As examples, deep brain stimulation (DBS) can be focused precisely but requires surgical implantation. In contrast, other popular neuromodulation methods include transcranial magnetic stimulation (TMS), electroconvulsive therapy (ECT), and transcranial direct current stimulation (tDCS). These techniques are non-invasive, but have reduced spatial resolution, and are further unable to target subcortical structures without also stimulating nearby regions. Unlike other methods, the emerging technology of focused ultrasound offers the unique possibility of targeted neuromodulation non-invasively. However, the safety and feasibility of this technology has yet to be tested adequately in human populations.

Low-Intensity Focused Ultrasound Pulsations (LIFUP) is a promising new potential neuromodulation tool, because it can target almost any region in the brain, including deep structures, with high spatial specificity and minimal effect on other regions^1^. LIFUP allows for the non-invasive delivery of acoustic energy to a well-localized and circumscribed brain region of a few millimeters in diameter, depositing mechanical or thermal energies^2,3^.

The administration of low-intensity focused ultrasound pulsation (LIFUP) has been demonstrated to a have a variety of neuromodulatory effects. These effects were non-thermal and reversible^4^. Further, the short acoustic wavelengths of high frequency ultrasound enable focusing the sonication to regions limited to several millimeters and, by increasing the surface area of the scalp over which the ultrasound is applied (either with larger spherical-section transducers, or using phased arrays), the focal spot can be placed essentially anywhere within the brain^2^ while depositing minimal energy outside of the targeted region. LIFUP differs from high-intensity focused ultrasound (HIFU) because the LIFUP energies are an order of magnitude lower than HIFU. Whereas HIFU is administered continuously for ablation, LIFUP is administered in short pulses, which reduces total energy deposition. Given these advantages, LIFUP is a tool with great potential in both therapy and diagnostics^5^.

Early investigations assessed safety with various ex vivo preparations. Repeated stimulation of brain slices every 8 minutes for 36 hours did not result in changes to the cytoarchitecture or integrity^6^. Upon further examination, there was no evidence of damage to the integrity of the BBB, and there was not a difference in the frequency of apoptotic neurons^7^. Additional investigations have been conducted using a variety of histologic assessments, using hematoxylin and eosin (H&E) staining, Terminal deoxynucleotidyl transferase dUTP nick end labeling (TUNEL), and vanadium acid fuchsin (VAF) staining with toluene blue counterstaining, among others, all of which did not show damage^8-11^.

A study of sheep brain showed microhemorrhaging after repetitive sonications of V1 using spatial peak, pulse average intensity (I_sppa_) of 6.6 W/cm^2^ with a 50% duty cycle^12^. It is worth noting that edema, cell necrosis, or localized inflammatory processes were not detected with the H&E staining that was used. Indeed, reported adverse effects from LIFUP are exceedingly rare^13^. There is evidence to suggest that at high intensity focused ultrasound stimulation, there may be effects such as hemorrhage, apoptosis, or opening of the blood-brain barrier (BBB). There may also be long-term changes in activity throughout the brain. It is however somewhat difficult to establish the safety of the technique, as published studies do not report parameters in a consistent manner.

The objective in the current study was to assess the safety and feasibility LIFUP in humans. To determine if LIFUP damages brain tissue, we utilized human participants with medication resistant temporal-lobe epilepsy who were already scheduled to undergo resective brain surgery for epilepsy treatment. This allowed application of LIFUP to the temporal lobe prior to its scheduled removal and then the detailed evaluation of the LIFUP-exposed tissue for histological damage.

This study provides preliminary evidence of safety and efficacy, which was evaluated using functional MRI (fMRI). Identifying LIFUP changes to the fMRI’s blood oxygenation level dependent (BOLD) signal provides an opportunity to assess for neuromodulation and therefore helps support the feasibility of LIFUP as a neuromodulatory treatment. The safety and efficacy of LIFUP was also assessed with electroencephalograms and neuropsychological testing immediately before and after the LIFUP exposure.

## Materials & Methods

All experimental procedures were approved by UCLA Institutional Review Board (IRB: 13-000670) and were regulated by an Investigational Device Exemption (IDE) G130290 from the US Food and Drug Administration (FDA).

### Participants

The participants were eight patients from the UCLA Seizure Disorders Center who were scheduled for an anterior-mesial temporal lobe resection as treatment for medication-resistant temporal lobe epilepsy. As part of the first-in-human safety testing, initial delivery of LIFUP was limited to 720 mW/cm^2^. Other small methodological and technical differences also existed in the first two participants. For these reasons, the first two participants are excluded from the analysis presented here.

The FDA initially limited inclusion to only those participants who were undergoing resection of the non-dominant hemisphere but later expanded subsequently the criteria to include both dominant and non-dominant temporal lobes. Criteria for inclusion were: age 18 - 60 years, epilepsy that has been resistant to at least 3 appropriate and FDA-approved anti-seizure medications, adherence to anti-seizure medication treatment, maintenance of a seizure diary, a seizure frequency of at least 3 seizures/month and an epilepsy surgery evaluation that identified unilateral hippocampal dysfunction and seizure onsets. The diagnostic evaluation included the intracarotid amobarbital procedure (IAP) and neuropsychological testing.

Participants who had a cognitive or psychiatric disorder that limited their ability to give informed consent were excluded. In the interest of safety, participants were excluded if the recent history included status epilepticus or seizures due to alcohol or illicit drugs. In addition, participants with implants or other metal components not compatible with MRI were excluded due to elevated risk. Lastly, pregnant participants were excluded. Participant demographic information is listed in table 1.

**Table 1.** Participant Demographics

| Participant | Age/Gender | Race | Side of Resection |
| --- | --- | --- | --- |
| 1 | 44/F | Hispanic | Right |
| 2 | 22/F | Hispanic | Right |
| 3 | 41/M | White/not Hispanic | Left |
| 4 | 24/F | Hispanic | Left |
| 5 | 27/M | White/not Hispanic | Left |
| 6 | 37/F | Asian/not Hispanic | Left |
| 7 | 25/M | White/not Hispanic | Right |
| 8 | 65/F | White/not Hispanic | Left |

### LIFUP Device

The study utilized various models of the BX Pulsar device (BrainSonix Inc., Sherman Oaks, CA). The BX Pulsar was designed to deliver LIFUP energy to the human brain, and to be safe for simultaneous use within a 3 Tesla MRI. The BX Pulsar transducer uses a spherically focused piezo element with a 6.1 cm aperture diameter and a fundamental resonance of 650 kHz. Acoustic intensity measurements in a water tank showed that the transducer has a focal maximum pressure at a depth of 6.2 cm and a focal volume (the region in which the pressure is more than one half the maximum pressure) that is a prolate sphere of approximately 4 mm × 4 mm × 28 mm in water.

For the first two participants, the transducer was placed within a water-filled holder with a membrane that allowed for deformation to fit the participant’s head surface. A second-generation transducer was used for the remainder of the study. The latter transducer incorporates an ultrasound conducting gel-pad (Pharmaceutical Innovations, Newark, NJ) between the transducer surface and the participant’s head. We applied ultrasound gel to both sides of the gel pad prior to putting the device onto the head, which provides the same contour benefit as the water-filled holder. This acoustic coupling is necessary to avoid impedance mismatches in the ultrasonic path and to ensure correct beam propagation and focus. For all participants, we attached the transducer was placed approximately over the temporal window and to the head with an adjustable strap.

### MRI and LIFUP Procedures

LIFUP was administered with MRI guidance to the temporal region on the side scheduled to undergo surgery, within the anterior temporal lobe. LIFUP was administered at least one day prior to the resection surgery. The ultrasound was focused on the region within the temporal lobe to be resected. Functional MRI of the brain was obtained throughout the LIFUP session.

Targeting of the anterior temporal lobe entailed an iterative process. We first placed the transducer in a position over the temporal bone. A brief 3D T1-weighted image was acquired to determine if the transducer was positioned accurately over the desired region. If not, the transducer was repositioned; we repeated this process as needed until we achieved proper placement. LIFUP was delivered concurrently during T2*-weighted BOLD imaging (TR=700 ms, TE=33ms, 2.5 mm^3^isotropic voxels, MB=6).

The LIFUP stimulus was administered under 2 different pulsing paradigms that were classified as “activation” and “suppression” based on previous preclinical research in animals. Activation LIFUP involved brief pulse trains with a 50% duty cycle. Suppression LIFUP involved 30-second pulse trains with a 5% duty cycle.

After our two initial participants received LIFUP at up to 720 mW/cm^2^ (I_spta_.3), and showed no evidence of tissue injury, all subsequent participants received the activation paradigm at four increasing intensities (I_spta_.3): 720 mW/cm^2^, 1440 mW/cm^2^, 2880 mW/cm^2^ and 5760 mW/cm^2^. For these participants, the activation paradigm was eight 0.5 sec bursts at 250 Hz PRF with a 0.2 msec pulse width and a 50% duty cycle. These participants also received the LIFUP suppression paradigm, which was identical to the previous participants, except it was administered at 5760 mW/cm^2^. One participant did not receive the suppression paradigm due to time constraints during the stimulation session.

### fMRI Analysis

FMRI data processing was carried out using FEAT (FMRI Expert Analysis Tool) Version 6.00, part of FSL (FMRIB’s Software Library, www.fmrib.ox.ac.uk/fsl). The following pre-statistics processing was applied; motion correction using MCFLIRT ^14^; non-brain removal using BET^15^; denoising using ICA-AROMA (automatic removal of motion artifact)^16^; spatial smoothing using a Gaussian kernel of FWHM 5mm; grand-mean intensity normalization of the entire 4D dataset by a single multiplicative factor; high-pass temporal filtering (Gaussian-weighted least-squares straight line fitting, with sigma=15.0s). BOLD data were analyzed with a general linear model approach including, as a regressor of interest, the onset and duration of each LIFUP sonication, convolved with a gamma response function to account for hemodynamic delay. Z (Gaussianised T/F) statistic images were thresholded using clusters determined by Z>2.3 and a (corrected) cluster significance threshold of P=0.05^17^.

### EEG

Electroencephalogram (EEG) was recorded immediately before and after the LIFUP/MRI procedures using an EGI 256 channel EEG (Electric Geodesics, Eugene, Oregon). Each EEG recording lasted approximately 30 minutes. The EEG recordings were reviewed by a neurologist certified in clinical neurophysiology who was blinded to the experimental setup.

### Neuropsychological Assessments

Before and after LIFUP/MRI procedures, participants underwent a neuropsychological battery to test the functioning of regions of the temporal lobe. The test battery was changed slightly as the study progressed in order to maximize internal consistency. The most used assessments were the Rey Auditory Verbal Learning Test (RAVLT), which evaluates verbal learning and memory, as well as either the Rey-Osterrieth Complex Figure Test (ROCFT), the Brief Visuospatial Memory Test-Revised (BVMT-R), or the Taylor Complex Figure Test (TCFT), all of which evaluate visuospatial learning and memory. It has been hypothesized that the RAVLT accesses predominantly dominant hemisphere functions^18^, whereas the other tests access predominantly non-dominant hemisphere functions^19^. For analysis of each test, “immediate recall” scores were used, and then converted to percentile ranks.

### Histology

After the scheduled surgical resection of the anterior-mesial temporal lobe, the removed tissue was fixed in 10% formaldehyde, processed for paraffin embedment and then sectioned at 10 µm. The slices were stained using terminal deoxynucleotidyl transferase dUTP Nick End Labeling (TUNEL), hematoxylin and eosin (H&E), and vanadium acid fuchsin (VAF) with toluidine blue counterstain. As a positive control, non-sonicated tissue also was processed and treated with DNase I to generate free 3’-OH ends. To visualize potential damage due to sonication, we stained adjacent paraffin sections with either with H&E, or VAF-toluidine blue. H&E staining is used customarily for examination of tissue integrity, whereas VAF-toluidine blue stained is performed to detect the presence of acidophilic neurons which are indicative of acute neuronal injury and subsequent apoptosis or necrosis. VAF staining also allows for the visualization of extravasation and blood vessel disruption^20^. Data was imaged at the UCLA Translational Pathology Core Laboratory (TPCL) using Applied Imaging Leica Aperio Versa.

## Results

We did not observe consistent BOLD signals across participants or ultrasound intensities. While we noted several suprathreshold signal increases, these activity clusters were noticed disproportionately outside of the targeted region and were distributed in locations including frontal, temporal, and parietal lobes. Seven participants had at least one fMRI with voxels that passed the cluster threshold for significance, but the significant voxel clusters were not related to the site of sonication nor to the LIFUP intensity (Figure 1).

**Figure 1.**
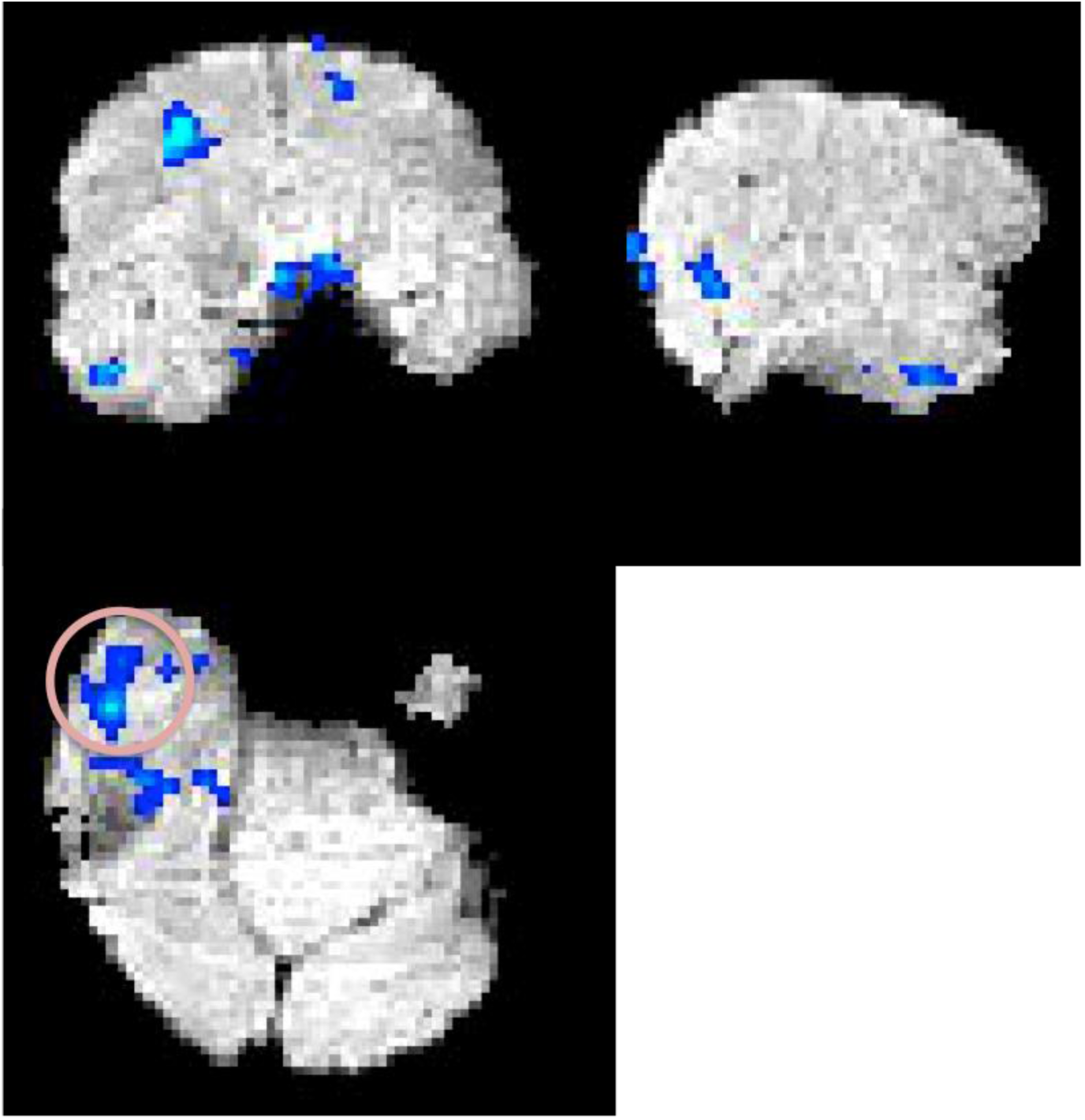
Example of LIFUP Suppression Paradigm This participant is exemplary of the overall results we found during the suppressive paradigm of either contralateral BOLD signal reduction or bilateral signal reduction. Including 3 orthogonal planes, this figure illustrates significant reductions in the BOLD signal corresponding to the LIFUP suppression paradigm. We have circled the significant temporal lobe cluster contralateral to the transducer. The transducer is not visible in this image.

For the LIFUP suppression paradigms a clearer picture emerged but it was not entirely consistent, picture emerged. Of the participants who received suppressive LIFUP at 5760 mW/cm^2^, 4 of 5 showed a cluster of significant BOLD signal reduction in the temporal lobe. For two participants the significant cluster was contralateral to the targeted hemisphere, while the other two participants showed significant clusters bilaterally.

Due to recording artifact and/or loss of data, a complete pre- and post- EEG was only available for 3 of the 8 participants; there was not sufficient data to perform a quantitative analysis. The EEG was unaffected by the stimulation procedure, but one participant’s EEG demonstrated slowing in the mid-temporal region after suppressive LIFUP, which indicates a functional effect.

The purpose of the neuropsychological testing was to determine whether the participants had any significant decay in cognitive capacity between pretest and posttest as a result of undergoing the LIFUP procedure, as assessed by the RAVLT and the BVMT-R (including, for some participants, either the ROCFT or the TCFT). A total of 8 participants were tested. They were divided into three groups. Participants in Group A (BX01-BX02) were administered both excitatory and inhibitory stimulation paradigms at a maximum intensity of 720mw/cm^2^.

Participants in Group B (BX05-BX08) were administered both excitatory and inhibitory stimulation paradigms at a maximum intensity of 5760mw/cm^2^. The participant in Group C (BX07) was only administered an excitatory stimulation paradigm at a maximum intensity of 5760mw/cm^2^. Neither Group A nor Group C had a sufficient number of participants to perform statistical analysis, so we dropped them from further consideration. We note that one of the participants in Group A (BX-02) performed better on a measure that evaluates verbal learning and memory pretest-to-posttest; and both participants in Group A (BX-01 and BX- 02) performed better on measures that evaluate visuospatial memory pretest-to-posttest.

As the data did not meet tests of normality, we used the non-parametric Wilcoxon Signed Rank Test, a non-parametric test to test the null hypothesis that LIFUP did not produce behavioral changes. for comparing two dependent samples. Our two-tailed null hypothesis was that the median of differences between pretest and posttest scores was zero at α = 0.05. On the RAVLT for Group B, there was a significant but only slight decline in mean scores pretest-to-posttest (pretest mean = 0.562 versus posttest mean −0.496). There was not a significant difference between pretest and posttest scores or Group B on those tests that evaluated visuospatial memory. These results are displayed at Table 2.

**Table 2.** Neuropsychological Differences From Pre- to Post-LIFUP for Group B using the Wilcoxon Signed Rank Test

| Test | W | z | p | r |
| --- | --- | --- | --- | --- |
| RAVLT | 15 | 2.023 | 0.043 | 0.640 |
| Visuospatial<br>Tests | 26.5 | -0.209 | 0.835 | -0.066 |

Examination of the H&E and VAF staining of samples from seven participants did not detect significant tissue damage, including necrosis, vascular damage, inflammation, significant gliosis, white matter damage, acidophilic/ischemic neurons, or extravasation. In one participant, observed acidophilic neurons and extravasation in the sonicated tissue were evident, but the significance of this finding is not known. The primary versus artifactual (procedural) nature of this finding in this case could not be ascertained. These changes were also noted in the non-sonicated areas from this patient’s resection material processed by both the UCLA Neuropathology Service and the Neurosurgery UCLA Rare Epilepsies & Brain Disease Tissue Bank (REBDTB) using TUNEL, H&E, and VAF staining. Therefore, these findings were inconclusive of LIFUP related effect.

## Discussion

In this study, we examined the safety of using transcranial LIFUP to modulate temporal lobe brain activity in participants with temporal lobe epilepsy scheduled for an anterior temporal lobe resection.

The measures of functional change, which included fMRI, EEG, and neuropsychological testing, were exploratory in this first-in-human study. The number of participants was small because of safety concerns, so results may not be fully reliable.

The simultaneous LIFUP fMRI experiments did not show consistent effects of LIFUP. Other research has also shown inconsistent LIFUP effects on fMRI, with only half of participants exhibiting significant BOLD changes^21^. That participants showed widely varying patterns of activation for the activation paradigm, does not allow for a definite conclusion. These suprathreshold clusters. The fMRI results may represent noise or artifact, as these are more likely when analyzing individual scans, and group level analysis was impractical because of the varying locations of our ultrasound targets across individuals and some variability may also be caused by differences in participant skull thickness that would affect the actual intensity of LIFUP experienced by the tissue. These observations may represent real activity related to the LIFUP stimulation by virtue of connectivity to the target tissue but this could not be determined given the other study limitations.

For the LIFUP suppression paradigms, the majority of participants receiving an intensity of 5760 mW/cm^2^ showed significant reductions in BOLD signal in the temporal lobe either contralaterally or bilaterally. The suppression paradigm differed significantly from the activation paradigm both in duration of pulse trains (30 sec vs 0.5 sec respectively) and in duty cycle (5% vs 50%) respectively. The longer duration represents a considerably higher delivered total deposition of LIFUP energy. Similarly, the lower duty cycle meant that the instantaneous mechanical energy delivered to the tissue was approximately ten times higher in the suppression paradigm. Given the evidence that LIFUP exerts its effects through mechanical interaction with the tissue^22^, it is not surprising that our results were more consistent when using the paradigm delivering greater mechanical energy.

Many factors may have contributed to the lack of consistent effect on the BOLD signal. First, targeting may have been different and activated different networks. The anatomy of the targeted region (the anterior temporal lobe) varied from participant to participant, so the exact aiming of the LIFUP likely differed between participants. Second, variations in skull thickness would affect the intensity achieved at the target tissue. Third, the targeted regions potentially contained abnormal (i.e. epileptogenic tissue) and may have responded differently to LIFUP than normal tissue. Other studies found inconsistent BOLD response^21^.

Other factors such as medication may have also played a role. For one, all participants differed in their anti-seizure medication regimens. Some participants experienced claustrophobia and were administered a 2 mg tablet of diazepam to take prior to the MR-guided LIFUP administration, and this also could affect the BOLD signal and the post-LIFUP neuropsychological testing.

Nevertheless, the neuropsychological testing revealed a significant decrease in participant’s verbal memory. As there was no sham procedure, we cannot know definitively if this resulted from the LIFUP itself, or some other aspect of the procedures. While we did use alternative forms of the RAVLT, these were not designed to be used on the same day and therefore may have been contaminated. Furthermore, participants were visibly fatigued from all the procedures, which likely affected their performance on the post-test.

The primary intent of the study was the determination of possible histologic damage from LIFUP, and there was no evidence of significant histopathologic damage was present in 7 of the 8 participants on light microscopy.

## Limitations

The size of this study limits its generalizability to a large population, but the results do not indicate substantial risk of LIFUP producing damage to brain tissue. The other results have greater limitations. Whether LIFUP produces minor changes to EEG or neuropsychological function cannot be determined from this study, and its effect on fMRI appears inconsistent. There is a possibility of histological changes at the molecular level, but this was not studied. Control of seizures was not evaluated – because temporal lobe epilepsy is usually infrequent and therapeutic effect could not be assessed within this experimental design.

## Data Availability

The data that supports the findings of this study are available from the corresponding author, upon reasonable request. The requesting PI will need an MTA.

## Conflict of Interest

Dr. Korb is Vice-President of BrainSonix Corp and owns shares in the company. Dr. Bystritsky is the Founder and CEO of BrainSonix Corp. Other authors report no conflicts of interest.

## Funding

This study received funding from the BrainSonix Corporation and the Friedman Foundation.

## Notes

### Clinical Trial

NCT02151175

## References

1. Spivak NM, Kuhn TP. Variations in targeting techniques of focused ultrasound for use in neuromodulation. Brain Stimul. 2019;12(6):1595–1596. doi:10.1016/j.brs.2019.07.021

2. Darrow DP. Focused Ultrasound for Neuromodulation. Neurotherapeutics.2019;16(1):88–99. doi:10.1007/s13311-018-00691-3

3. Dalecki D. Mechanical bioeffects of ultrasound. Annu Rev Biomed Eng. 2004;6(1):229–248. doi:10.1146/annurev.bioeng.6.040803.140126

4. Spivak NM, Schafer ME, Bystritsky A. Reversible neuroinhibition does not require a thermal mechanism. Brain Stimul. 2020;13(1):262. doi:10.1016/j.brs.2019.09.007

5. Blackmore J, Shrivastava S, Sallet J, Butler CR, Cleveland RO. Ultrasound Neuromodulation: A Review of Results, Mechanisms and Safety. Ultrasound Med Biol. 2019;45(7):1509–1536. doi:10.1016/j.ultrasmedbio.2018.12.015

6. Tyler WJ, Tufail Y, Finsterwald M, et al. Remote excitation of neuronal circuits using low-intensity, low-frequency ultrasound. PLoS One. 2008;3(10):e3511. doi:10.1371/journal.pone.0003511

7. Tufail Y, Matyushov A, Baldwin N, et al. Transcranial pulsed ultrasound stimulates intact brain circuits. Neuron. 2010;66(5):681–694. doi:10.1016/j.neuron.2010.05.008

8. Dallapiazza RF, Timbie KF, Holmberg S, et al. Noninvasive neuromodulation and thalamic mapping with low-intensity focused ultrasound. J Neurosurg. 2018;128(3):875–884. doi:10.3171/2016.11.JNS16976

9. Yang PS, Kim H, Lee W, et al. Transcranial focused ultrasound to the thalamus is associated with reduced extracellular GABA levels in rats. Neuropsychobiology. 2012;65(3):153–160. doi:10.1159/000336001

10. Yoo SS, Bystritsky A, Lee JH, et al. Focused ultrasound modulates region-specific brain activity. Neuroimage. 2011;56(3):1267–1275. doi:10.1016/j.neuroimage.2011.02.058

11. Min BK, Yang PS, Bohlke M, et al. Focused Ultrasound Modulates the Level of Cortical Neurotransmitters: Potential as a New Functional Brain Mapping Technique. Int J Imag Syst Tech. 2011;21(2):232–240. doi:10.1002/ima.20284

12. Lee W, Lee SD, Park MY, et al. Image-Guided Focused Ultrasound-Mediated Regional Brain Stimulation in Sheep. Ultrasound Med Biol. 2016;42(2):459–470. doi:10.1016/j.ultrasmedbio.2015.10.001

13. Pasquinelli C, Hanson LG, Siebner HR, Lee HJ, Thielscher A. Safety of transcranial focused ultrasound stimulation: A systematic review of the state of knowledge from both human and animal studies. Brain Stimul. 2019;12(6):1367–1380. doi:10.1016/j.brs.2019.07.024

14. Jenkinson M, Bannister P, Brady M, Smith S. Improved Optimization for the Robust and Accurate Linear Registration and Motion Correction of Brain Images. Neuroimage. 2002;17(2):825–841. doi:10.1006/nimg.2002.1132

15. Smith SM. Fast robust automated brain extraction. Hum Brain Mapp. 2002;17(3):143–155. doi:10.1002/hbm.10062

16. Pruim RHR, Mennes M, van Rooij D, et al. ICA-AROMA: A robust ICA-based strategy for removing motion artifacts from fMRI data. Neuroimage. 2015;112:267–277. doi:10.1016/j.neuroimage.2015.02.064

17. Worsley KJ. Statistical analysis of activation images. In: Jezzard P, Matthews PM, Smith SM, eds. Functional MRI: an introduction to methods. New York: Oxford University Press Inc.; 2001:251–270.

18. Vakil E, Blachstein H, Wertman-Elad R, Greenstein Y. Verbal learning and memory as measured by the Rey-Auditory Verbal Learning Test: ADHD with and without learning disabilities. Child Neuropsychol. 2012;18(5):449–466. doi:10.1080/09297049.2011.613816

19. Bonner-Jackson A, Mahmoud S, Miller J, Banks SJ. Verbal and non-verbal memory and hippocampal volumes in a memory clinic population. Alzheimers Res Ther. 2015;7(1):61. doi:10.1186/s13195-015-0147-9

20. Victorov IV, Prass K, Dirnagl U. Improved selective, simple, and contrast staining of acidophilic neurons with vanadium acid fuchsin. Brain Res Brain Res Protoc. 2000;5(2):135–139.

21. Ai L, Mueller JK, Grant A, Eryaman Y, Legon W. Transcranial focused ultrasound for BOLD fMRI signal modulation in humans. 2016 IEEE 38th Annual International Conference of the Engineering in Medicine and Biology Society (EMBC); 2016.

22. Lemaire T, Neufeld E, Kuster N, Micera S. Understanding ultrasound neuromodulation using a computationally efficient and interpretable model of intramembrane cavitation. Journal of neural engineering. 2019;16(4):046007. doi:10.1088/1741-2552/ab1685

